# Superspreading and the evolution of virulence

**DOI:** 10.1101/2025.09.03.25335001

**Authors:** Xander O’Neill, Andy White, Graham R. Northrup, Chadi M. Saad-Roy, P. Signe White, Mike Boots

**Author notes:** Corresponding author: x.o’. X.O. and A.W. contributed equally to this work.

## Abstract

Superspreading, where a small proportion of a population can cause a high proportion of infection transmission, is well known to be important to the epidemiology of a wide range of pathogens, including SARS-CoV-2. However, despite its ubiquity in important human and animal pathogens, the impact of superspreading on the evolution of pathogen virulence is not well understood. Using theory and both deterministic and stochastic simulations we examine the evolution of pathogen virulence under a range of different distributions of infection transmission for the host. Importantly, for many pathogens, superpreader events may be associated with increased tolerance to infection or asymptomatic infection and when we account for this super-spreading selects for higher virulence. In contrast, in animal populations where highly connected individuals, that are linked to superspreader events, also have fitness benefits, superspreading may select for milder pathogens. In isolation, the transmission distribution of the host does not impact selection for pathogen virulence. However, superspreading reduces the rate of pathogen evolution and generates considerable variation in pathogen virulence. Therefore, the adaptation of an emerging infectious disease, that exhibits superspreading, is likely to be slowed and characterised by the maintenance of maladaptive variants. Taken as a whole, our results show that superspreading can have important impacts on the evolution of pathogens.

**Author Summary:** The impact of infectious disease can vary from individual to individual. Superspreader events, where a few individuals cause a high proportion of infections, are critical to the spread and outbreak size of a wide range of important infectious diseases of humans and animals. Superspreading events and pathogen evolution, were important features of the Covid-19 pandemic, highlighted by the succession of dominant strains during the pandemic. There is a clear need to understand how superspreader events will affect the evolution of pathogens, in particular how the level of virulence (the additional mortality due to the infection) will evolve. In this study we use mathematical models to show that superspreading reduces the rate of pathogen evolution and generates considerable variation in pathogen virulence. Therefore, the adaptation of an emerging infectious disease, that exhibits superspreading, is likely to be slowed and characterised by the maintenance of maladaptive variants. Importantly, for many pathogens, superpreader events may be associated with increased tolerance to infection or asymptomatic infection, and when we account for this, superspreading selects for higher virulence. Our results show that superspreading can have important impacts on the evolution of pathogens.

## Introduction

Recent epidemics have emphasised the importance of infectious disease to human health, agricultural and natural wildlife systems [1–4]. It is now recognised that the management of epidemic and endemic infection requires well-developed theory, not only on the ecology/epidemiology, but also on the evolution of pathogens [5, 6]. The SARS-CoV-2 epidemic has further emphasised a key feature of many important pathogens, that there is considerable heterogeneity between individuals in infection transmission levels and disease severity [7–9]. In particular, superspreader events, where some individuals infect a considerably higher proportion of the population than an average individual [3, 7, 10, 11], are critical to the epidemic outcome. As the proportion of the population that are superspreaders increases, both the likelihood of disease extinction and size of infectious outbreaks also increase [10–14]. In the context of evolution it has been shown that superspreading can suppress the invasion of new strains and may slow disease adaptation [13, 14]. However, the impact of superspreading on the evolution of virulence (defined throughout as the increase in mortality due to infection) is unclear and given both the ubiquity of superspreading and the central role of the evolution of pathogens to their epidemiological impact this is an important knowledge gap [12].

Understanding the evolution of pathogen virulence and transmission is important for understand-ing emergent disease outcomes, for the development of disease management strategies, and to assess the burden of infectious disease in natural populations [15, 16]. Much of our current knowledge is based on the results of theoretical studies that make predictions on how virulence may evolve under a wide range of conditions [15]. This well-developed theory commonly assumes a saturating trade-off between transmission and virulence and makes general predictions [17–19], including how an increase in host mortality (and therefore a reduction in host lifespan) selects for increased transmission and virulence [15, 18], a result that holds when the host mortality increase is due to predation [20], the host immune response [21], or culling [22]. There is also theory on the impact of host population structure on the evolution of virulence, showing that as spatial contact structure changes, from global to highly local, virulence typically evolves to a lower level [23–26].

The effect of host heterogeneity on the evolution of pathogen virulence has been analysed in systems that consider two distinct host types [27, 28], when infection control can partition a single host based on infection risk [15, 29], and through control with imperfect vaccines [30, 31]. When a pathogen can infect two host types and pathogen virulence in each host was linked via a trade-off, theory has shown that pathogen evolution in a heterogeneous host population may evolve to specialise for one of the two host types at the cost of losing its specialisation for the other type [27]. With infection control, host heterogeneity can have an impact on pathogen evolution when vaccination is targeted at a more vulnerable subpopulation. Transmission then becomes concentrated on the unvaccinated subpopulation which can change the evolved level of virulence compared to uniform vaccine coverage [15, 29]. Similarly imperfect vaccines lead to heterogeneity between vaccinated and unvaccinated hosts and can select for an increase or decrease in virulence depending on the pathogen mechanisms that the vaccine targets [30, 31]. There is also theory on the coevolution of sociality and pathogen virulence that assumes variation across hosts in contacts and transmission [32, 33], but these studies do not assess pathogen evolution for different contact distributions. Importantly, the variation between individuals in terms of infection transmission is often related to other disease characteristics [34, 35], in particular, superpreader events may be associated with increased tolerance to infection or asymptomatic infection, as observed in the COVID-19 pandemic [7–9]. As such, while previous studies have highlighted that various forms of host heterogeneity can play a role in determining virulence evolution, a clear focus on how superspreading will affect the evolution of virulence is still required.

Previous work has shown that superspreading events can suppress the emergence of new pathogen strains [13] and that superspreading, represented as a change in the number of host contacts, has no effect on virulence evolution in classical *SI* models [36]. However, the impact on virulence of a range of contact/transmission structures, that characterise increasing levels of superspreading, has not been examined in detail. Moreover, transmission heterogeneity and in particular, superspreading, may be associated with individual level variation in other host characteristics including host natural mortality, increased disease-induced mortality (vulnerability), or decreased disease-induced mortality (tolerance) [34, 35], and the impact of these associations has not been explored. Using both deterministic and stochastic model frameworks, we address these knowledge gaps by examining the impact of a range of contact/transmission structures and how the association between superspreading and other host and pathogen characteristics impact the evolution of virulence. In line with previous theoretical studies that examine pathogen evolution (see [15]), we will develop a general model framework for an endemic infectious disease system and use this to test and explain how changing from a homogeneous to an increasingly heterogeneous infection transmission distribution for the host will effect pathogen selection for virulence. Our results show how superspreading impacts the rate of evolution, the diversity of evolved pathogen strains, the persistence of maladaptive variants, and importantly the evolved level of pathogen virulence and transmission.

## Methods

In a seminal paper on the epidemiological impacts of superspreading, Lloyd-Smith et al. (2005) [11] assumed variation in infection transmission between individuals and confronted models with data for a wide range of important human infectious diseases. They showed how superspreading can be captured by a gamma distribution that represented the individual level variation in the number of secondary cases of infection. We follow this established approach and represent heterogeneity by a gamma distribution that represents variation in infection transmission across host individuals [11]. The shape parameter for the gamma distribution governs the transmission distribution for the host (Figure 1), which ranges from homogeneous (all host individuals have the same level of transmission), to superspreading (a few host individuals have a high level of transmission, and most individuals have a low level of transmission). Importantly, we assume that the different transmission distributions have the same mean level of transmission.

**Figure 1:**
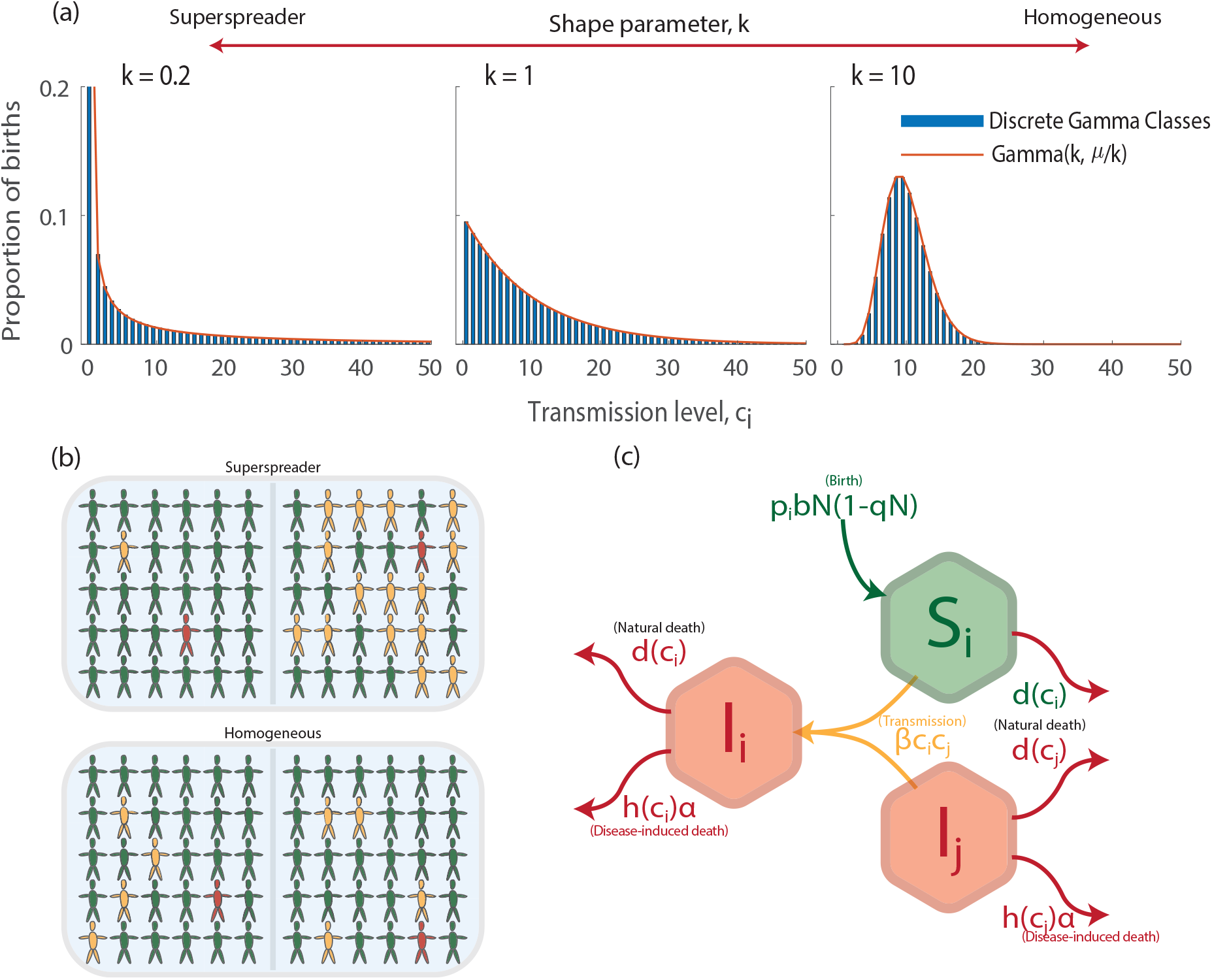
(a) The distribution of host transmission levels for different shape parameters of the gamma distribution, *k*, showing the probability of hosts being born, *p*_*i*_, with a particular level of transmission, *c*_*i*_. As *k* increases the transmission distribution transitions from superspreading to homogeneous. The red line shows the exact gamma distribution and the blue bars our discretised version used in simulations. The mean transmission level is the same in all distributions. In (b) we highlight possible transmission events from an infected individual (red) that can infect individuals (yellow) from a pool of susceptible individuals (green). We capture superspreading, where an individual may infect few individuals (top left) or many individuals (top right), and a homogeneous transmission distribution where an infected individual always infects the same number of susceptible individuals (bottom left and right). In (c) we show a schematic of our model (Equations 1) highlighting how infection from an infected of type *I*_*j*_ of a susceptible of type *S*_*i*_ leads to an infected of type *I*_*i*_.

To assess the impact of superspreading on pathogen evolution we extend a classical susceptible-infected (*SI*) epidemiological model [37, 38] where a susceptible, *S*, can become infected, *I*, through direct contact. The susceptible and infected classes are partitioned into *n*_*c*_ host types based on their transmission level, with an individual of host type *i* having transmission level *c*_*i*_. The model for the population density of individuals of host type *i* is as follows:

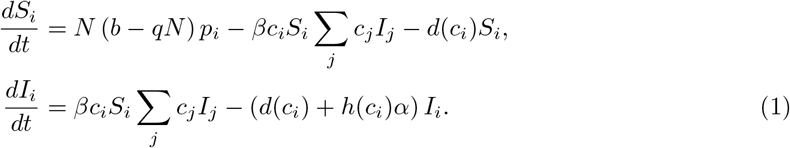

Here, *N* = Σ_*i*_ *S*_*i*_ + Σ_*i*_ *I*_*i*_ denotes the total population density, *b* denotes the maximum birth rate, and *d*(*c*_*i*_) denotes the natural, non-disease related, death rate for type *i* (with transmission level *c*_*i*_). Note, we initially assume *d*(*c*_*i*_) = *d* and so is independent of the transmission level of the host. Later we relax this assumption. The parameter *q* represents the population’s susceptibility to crowding and is set such that the population size is equal to its carrying capacity, *N* = *N*_*K*_, in the absence of infection. When *d*(*c*_*i*_) = *d* this implies *q* = (*b − d*)*/N*_*K*_. When *d*(*c*_*i*_) = *d*_*i*_ varies with *c*_*i*_ then *q* = (*b −* 1*/* Σ_*i*_ (*p*_*i*_*/d*_*i*_)) */N*_*K*_.

We assume a proportion, *p*_*i*_, of all births occur into susceptible class, *S*_*i*_. This proportion follows a truncated, discrete, gamma probability distribution function with shape parameter *k* and mean 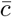, following the scale-shape definition of a gamma distribution, Γ(*k, θ*), where 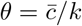. Here 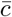 is the average level of transmission for the host and this remains fixed for all transmission distributions. We truncate the probability distribution between 0 and 100, resulting in *n*_*c*_ = 100 classes. We discretise the range [0, 100] into unit intervals so that we can have finite classes in the models. Each *c*_*i*_ takes the expected value of the probability distribution within the respective unit interval. We consider a range of gamma distributions, with the restriction that 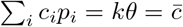, and where the transmission distribution for the host can range from homogeneous (large *k*), to heterogeneous (small *k*). This framework characterises distributions with an increasing probability of superspreading as *k* decreases (Figure 1).

We assume density-dependent infection transmission with the transmission coefficient for an infected individual of type *j* infecting a susceptible individual of type *i* given by *βc*_*i*_*c*_*j*_. Here *β* is the transmission component controlled by the pathogen, the transmission level *c*_*i*_ equates to the susceptibility of host type *i*, and *c*_*j*_ to the infectivity of host type *j*. Our default assumption is that a superspreader will have high susceptibility and high infectivity, and this emerges naturally when heterogeneity in transmission occurs through host contacts. However, other mechanisms that lead to heterogeneity in transmission in the host, particularly those that have a biological basis, may impact either susceptibility or infectivity only and it is critical to distinguish between superspreading as a consequence of social behaviour/contact rate heterogeneity, and superspreading rooted in biological factors [34]. We account for this by adjusting the transmission coefficient to 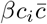 if there is heterogeneity in host susceptibility only, and to 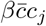 if it impacts infectivity only. This set-up allows us to consider different underlying mechanisms that could lead to heterogeneity in *c*_*i*_, such as behavioural mechanisms that lead to variation in host contacts or physiological mechanisms that may lead to variation in host vulnerability or tolerance [34, 35] and highlights the flexibility of the transmission term to capture several different aspects of superspreading.

Infected individuals incur disease-induced mortality (virulence) at rate *h*(*c*_*i*_)*α*. Here, *α* is the contribution to virulence set by the pathogen. We initially assume the host component of virulence is independent of host type *h*(*c*_*i*_) = 1. If we instead assume that host infection transmission is related to host vulnerability, then hosts with high susceptibility to infection incur additional disease induced mortality, represented by taking *h*(*c*_*i*_) to be an increasing function of *c*_*i*_. If hosts with high infectivity are more tolerant of the infection, then *h*(*c*_*i*_) is a decreasing function of *c*_*i*_ [39–42]. Parameter descriptions are detailed in Table 1.

**Table 1:** Demographic and epidemiological model parameter descriptions and baseline values.

| Parameter | Description |
| --- | --- |
| $N_K = 1000$ | Carrying capacity of the population. |
| $b = 10$ | Maximum birth rate of a susceptible. |
| $d(c_i)$ | Natural death rate for hosts of type $i$ ($d(\bar{c}) = d = 1$). |
| $q(c_i)$ | Susceptibility to crowding ($q(\bar{c}) = 0.009$). |
| $h(c_i)$ | Host contribution to virulence ($h(\bar{c}) = 1$). |
| $n_c = 100$ | Number of transmission classes for the host. |
| $c_i \in [0, 100]$ | Infection transmission level for hosts of type $i$ ($\bar{c} = 10$). |
| $p_i$ | Proportion of births for hosts of type $i$. |
| $n_\alpha = 101$ | Number of pathogen virulence/transmission classes. |
| $\beta \in [\beta_{min}, \beta_{max}]$ | Pathogen controlled transmission coefficient. |
| $\beta_{min} = 0.00002$ | |
| $\beta_{max} = 0.001$ | |
| $\alpha \in [\alpha_{min}, \alpha_{max}]$ | Pathogen controlled virulence level. |
| $\alpha_{min} = 0$ | |
| $\alpha_{max} = 10$ | |
| $a = -0.25$ | Curvature of the transmission/virulence trade-off function. |

### Evolution of virulence

This model framework (Equations 1) can be used to simulate the epidemiological dynamics for different transmission distributions (determined by *k*). In this study we will examine how pathogen virulence will evolve under a range of different distributions for infection transmission of the host (that is, for each *k* we examine how *α* evolves). We utilise three different modelling techniques to model the evolutionary process: adaptive dynamics [43, 44], deterministic simulations, and stochastic simulations. We use adaptive dynamics to provide analytical expressions for the evolutionarily singular strategy (ESS) for virulence. We use this approach specifically when *d*(*c*_*i*_) = *d* and *h*(*c*_*i*_) = 1, as here analytic expressions are tractable. We use deterministic simulations to illustrate the analytical findings, and to determine ESS virulence for scenarios where the adaptive dynamics analysis is intractable (when *d*(*c*_*i*_) and *h*(*c*_*i*_) are not constant). We use stochastic simulations to provide information on the rate of evolution to the ESS and the variation in virulence.

All three modelling techniques include a mutation process that allows new strains of the pathogen (with different values of *α*) to emerge and potentially replace the current, resident, pathogen strain. Throughout, and in line with previous studies [15, 17, 18], we assume that there is a trade-off between *β* and *α* such that benefits to the pathogen in terms of increased transmission are bought at a cost of increased virulence [17–19]. Mathematically this means *β* = *f* (*α*) where *f′*(*α*) *>* 0 and *f′′*(*α*) *<* 0 [45]. This ensures virulence evolves to an evolutionarily stable strategy (ESS). For the deterministic and stochastic simulations we require an explicit expression for the trade-off between transmission and virulence [22, 46]. This takes the following form:

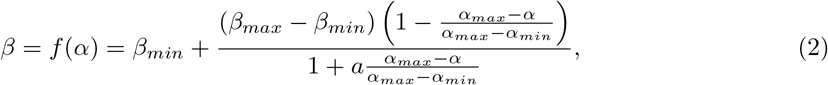

where *β*_*min*_, *β*_*max*_ denote the minimum and maximum values for *β*, respectively, *α*_*min*_, *α*_*max*_ denote the minimum and maximum values for *α*, respectively, and *a* represents the curvature of the trade-off function (see Figure S.1).

### Adaptive dynamics

Adaptive dynamics assumes a separation of epidemiological and evolutionary time scales such that the epidemiological dynamics have reached a dynamic attractor before mutation of a new pathogen strain is considered [43, 44]. When mutation occurs, a mutant strain, with small phenotypic variation from the resident strain, emerges (at low density) and in our study competes with the established resident strain that is at its endemic steady state. The success of the mutant strain depends on its fitness (its long-term growth rate). If the fitness of the mutant strain is negative it will die out. If the fitness of the mutant strain is positive the mutant strain can invade and replace the resident strain, to become the resident itself. Adaptive dynamics considers multiple steps of this mutation and replacement process until the pathogen converges to an ESS.

### Deterministic Simulations

To simulate the adaptive dynamics process we split the infected classes into *n*_*α*_ = 101 pathogen strains based on their virulence (and transmission), with strains split uniformly across the interval [*α*_*min*_, *α*_*max*_]. A strain *z*, has virulence, *α*_*z*_ and pathogen transmission factor *β*_*z*_ = *f* (*α*_*z*_). This requires extending Equations (1) to include *n*_*α*_ pathogen strains that include the density of hosts of type *i* that are infected with pathogen strain *z*. The extended model is as follows:

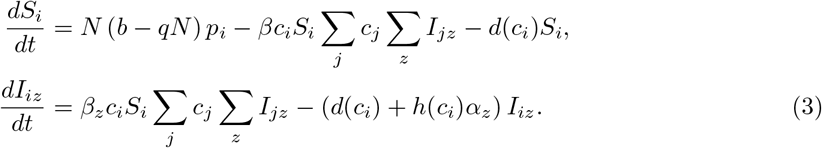

We use the parameter values stated in Table 1 which are a general parameter set chosen to ensure the system exhibits a stable, positive, endemic steady state for all pathogen strains. We select an arbitrary initial pathogen strain, *z* = 31 (and so *α*_*z*_ = 3 with corresponding *β*_*z*_ = 0.00038) and initial total densities of *S* = 151 and *I* = 584 (the endemic steady state densities when *α* = 3), which are split for each host class *i* based on probabilities *p*_*i*_. We then numerically solve Equations for a fixed time, *t*_*e*_. This allows the epidemiological dynamics to approach the endemic steady state for that particular pathogen virulence/transmission level. A mutant pathogen strain is then introduced, at low infected density, with a pathogen virulence level close to the current resident strain (the mutant strain is selected as either the strain directly above or below the resident strain, with equal probability). The population dynamics are numerically solved for a further time, *t*_*e*_, to allow the epidemiological dynamics to approach the steady state, where either the mutant strain dies out or the mutant replaces the resident strain. This procedure is then repeated, and allows the pathogen to evolve to the ESS, *α*^*∗*^. These simulation methods have been successfully used to approximate the adaptive dynamics process [43,45], but it should be noted that in this approximation the epidemiological dynamics may not necessarily reach their steady state before a new mutation arises. In this way, the ecological and evolutionary time scales are not strictly separated, as assumed in adaptive dynamics theory.

### Stochastic Simulations

We relax the assumptions of adaptive dynamics by considering a stochastic, individual based model of Equations (3) where susceptible, *S*, and infected, *I*, populations levels take integer values. As in the deterministic simulations, we use a trade-off between pathogen virulence and transmission (see Equation 2), the parameter values outlined in Table (1), and our default assumption on heterogeneous transmission (that an individual with high susceptibility would also have high infectivity). The key differences between the stochastic simulations and the deterministic simulations are that births are now individual events that occur with probability *p*_*i*_ to a specific host type (rather than proportionately to all host types), and that mutations now occur with a small probability, *ϵ* = 0.002, whenever an infection event occurs (rather than once the population has approached its endemic steady state). The mutant strain is selected as either the strain directly above or below the strain of the infecting individual, with equal probability. Simulations were undertaken using a Gillespie algorithm [47,48] (continuous-time Markov chain), where a specific individual event occurs at random according to the relative transition rates for each event (see Table 2). The population classes and transition rates are updated after each event, and the time between events was taken from an exponential distribution with rate equal to the sum of the transition rates. Notably, the probabilistic and individual based nature of these simulations can result in the stochastic fade-out of pathogen strains.

**Table 2:** Transition events and their respective rates for the stochastic model framework. In addition, we assume that infection can lead to mutation to a neighbouring pathogen strain with probability *ϵ* = 0.002. To determine all the possible transition events, it is necessary to calculate the transition rates for all host types *i* and for all possible infection interactions for host type *i*.

| Transition | Transition rate | Description |
| --- | --- | --- |
| $S_i \rightarrow S_i + 1$ | $N(b - qN)p_i$ | Birth of a susceptible of host type, $i$. |
| $S_i \rightarrow S_i - 1$ | $d(c_i)S_i$ | Natural death of a susceptible of host type, $i$. |
| $S_i \rightarrow S_i - 1,$<br>$I_{iz} \rightarrow I_{iz} + 1$ | $\beta_z c_i S_i c_j I_{jz}$ | Transmission of infection from an infected individual of host type, $j$, with strain $z$ of the pathogen ($I_{jz}$) to a susceptible individual of host type, $i$. |
| $I_{iz} \rightarrow I_{iz} - 1$ | $(d(c_i) + h(c_i)\alpha_z)I_{iz}$ | Death of an infected of host type, $i$, with strain $z$ of the pathogen. |

## Results

### Transmission level of the host is independent of other host characteristics

In our model set-up infection transmission of the host is independent of other host characteristics when *h*(*c*_*i*_) = 1 and *d*(*c*_*i*_) = *d*. Using adaptive dynamics we assess how virulence will evolve by determining the fitness of a mutant strain of a pathogen (with parameter *α*_*M*_). We determine conditions that allow the mutant pathogen strain to invade a population with a resident, endemic, pathogen strain (with parameter *α*_*R*_). In line with Van Baalen (2002) [36] it can be shown that the fitness of the mutant strain is positive if the following condition is satisfied (see supplementary material, Section S.1, for further details):

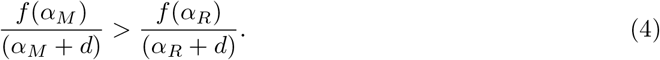

Therefore, any mutant strain that satisfies Equation (4) will replace the resident strain and the pathogen evolves a level of virulence, *α*^*∗*^, that maximizes *f* (*α*)*/*(*α* + *d*), which is the optimal strategy [49]. This is independent of the transmission level of the host (independent of *c*_*i*_) and so when *h*(*c*_*i*_) = 1 and *d*(*c*_*i*_) = *d* pathogen virulence will evolve to an ESS at *α*^*∗*^ for all transmission distributions of the host. Note, under the same assumptions, other model structures (*SIS, SIR, SIRS*) also lead to the same finding that *α*^*∗*^ is independent of the transmission level of the host. We show the analysis for the *SIRS* model structure with either density-dependent or frequency-dependent transmission in the supplementary material (Section S.1) and note that the *SIRS* model framework encompasses the other model structures.

We use deterministic simulations to illustrate the adaptive dynamics results. The deterministic simulations indicate that the pathogen evolves to the same level of virulence *α*^*∗*^ for different transmission distributions for the host (Figure 2a). This confirms the adaptive dynamics analysis and indicates that the deterministic simulations provide a robust method for determining ESS virulence.

**Figure 2:**
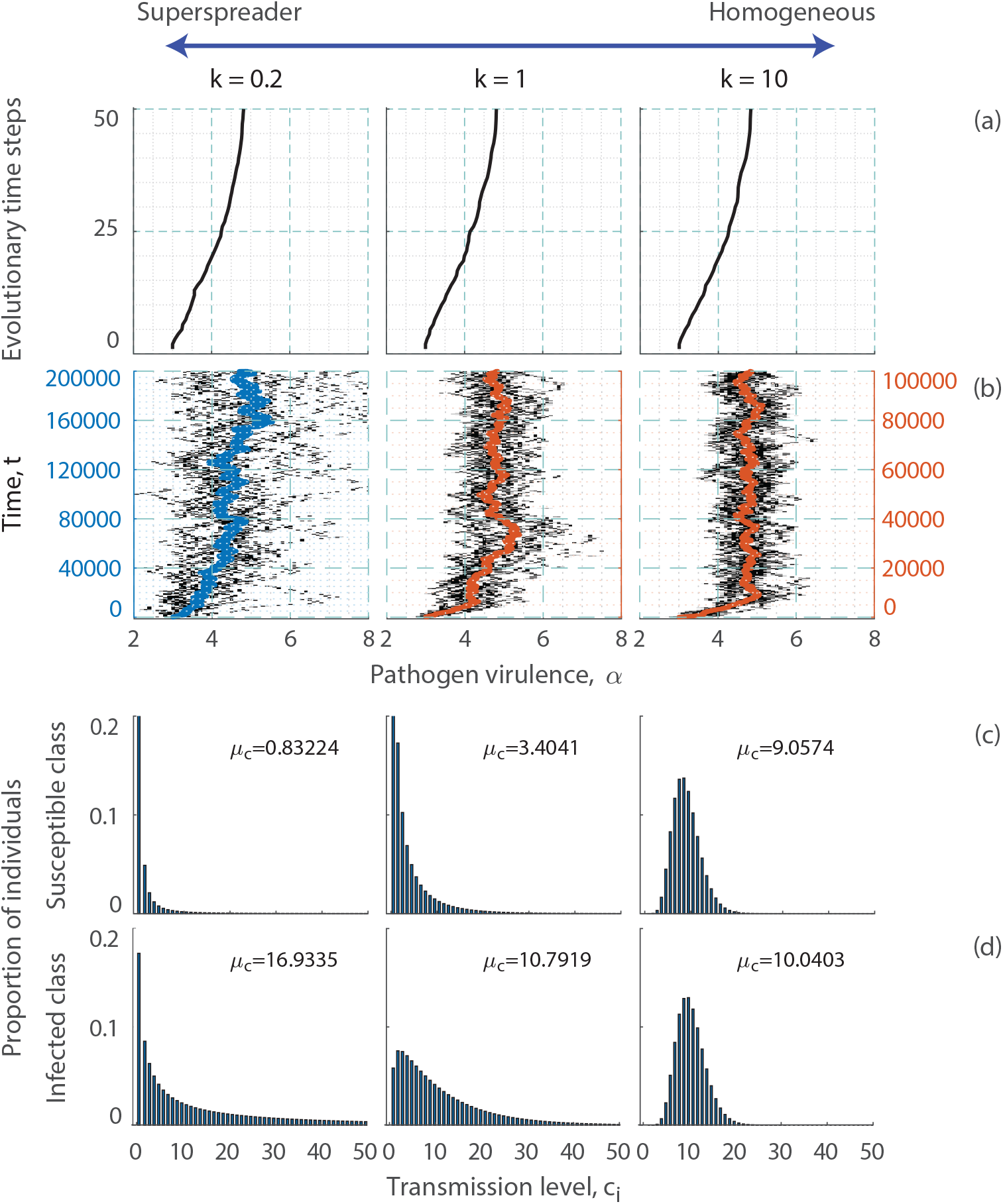
The effect of superspreading on the evolution of virulence for the *SI* model when infection transmission for the host is independent of other host characteristics. In (a) and (b) we show the evolution of pathogen virulence over time under different transmission distributions. In (a) we show the deterministic simulations and (b) we show the stochastic simulations (and note the vertical axis is different for *k* = 0.2 compared to *k* = 1 and *k* = 10). In (c) we show the proportion of susceptible individuals in each transmission class, *c*_*i*_ and (d) the proportion of infected individuals in each transmission class. All proportions are shown at the evolutionary stable level of pathogen virulence, *α*^*∗*^, in the deterministic simulations. The mean level of transmission, *µ*_*c*_, is also shown for each distribution. The variance in *α* over the last 1000 time points of the stochastic simulations is as follows: *k* = 0.2, variance = 1.6; *k* = 1, variance = 0.34; *k* = 10, variance = 0.23. Parameters are taken from Table 1.

### Rate of evolution and variability in virulence

Under the stochastic framework, we confirm previous results, that pathogen virulence evolves to *α*^*∗*^ and that this is not affected by the transmission distribution of the host (Figure 2b). However, the rate at which virulence evolves and the variability in virulence is different for different transmission distributions (Figure 2b). In particular, for heterogeneous transmission distributions that can represent superspreading, the rate of evolution to *α*^*∗*^ is slowed and the variability in virulence is increased. Under superspreading, the proportion of hosts with high transmission is small, the infected population has a higher proportion of hosts that had high susceptibility and have high infectivity, than the susceptible population that has a higher proportion of hosts with low susceptibility (see Figure 2c and 2d and the reported average level of transmission of the host, *µ*_*c*_). Since mutation occurs upon infection of a susceptible host, the mutation is likely to reside in a host that had low susceptibility and therefore low infectivity. This means the mutation either spreads slowly or fades-out due to stochastic effects, and explains why the rate is reduced and variability is increased when progressing towards the evolutionarily stable level of virulence, *α*^*∗*^. Therefore, our first key result is that superspreading *per se* does not impact the long-term evolution of pathogen virulence, but it slows adaptation and generates diversity.

We find that the increase in variation in virulence and reduction in the rate of adaptation as heterogeneity increases holds when we consider different transmission terms that depend on susceptibility and infectivity, susceptibility only, or infectivity only (Figure S.2). When heterogeneity applies to infectivity only (a transmission term of 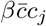) then all individuals are equally susceptible, but only a few of the individuals that become infected will have high infectivity and so most mutations occur in individuals with low infectivity. This slows the rate of evolution to *α*^*∗*^ and increases variability in virulence compared to the case with homogeneous transmission (Figure S.2b and S.2d). When heterogeneity applies to susceptibility only 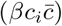 then individuals with higher susceptibility are more likely to become infected and therefore the remaining susceptible population will have reduced (less than average, 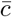) susceptibility. This reduced susceptibility means that when a mutation occurs upon infection the infected individual with the mutant strain will have a reduced chance of causing onward infection, compared to when there is a homogeneous transmission distribution. This, again, reduces the rate of evolution to *α*_*∗*_ and increases variability in virulence (Figure S.2c and S.2d).

### Transmission level of the host is correlated with other host characteristics

We also consider scenarios where infection transmission is associated with other host characteristics, including host lifespan, tolerance to infection, and vulnerability to infection. Whilst determining the fitness of the mutant strains is possible, the analytical expressions for the fitness function and for ESS virulence are complex and do not provide tractable information. Therefore, we find ESS virulence using deterministic simulations and stochastic simulations.

#### High contact levels increase host lifespan

Individuals with many contacts may gain fitness benefits, exemplified by the observation that an animals sociability can increase access to survival-related information [50, 51]. This increased access can then lead to a reduction in non-disease related mortality (in line with [32, 33]). To represent this, we assume *c*_*i*_ represents the contact level for hosts of type *i* and that *d*(*c*_*i*_) is a decreasing function of *c*_*i*_, with 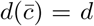 (Figure 3b). In this scenario, we find pathogen virulence decreases as the contact distribution for the host changes from homogeneous to superspreading (Figure 3a).

**Figure 3:**
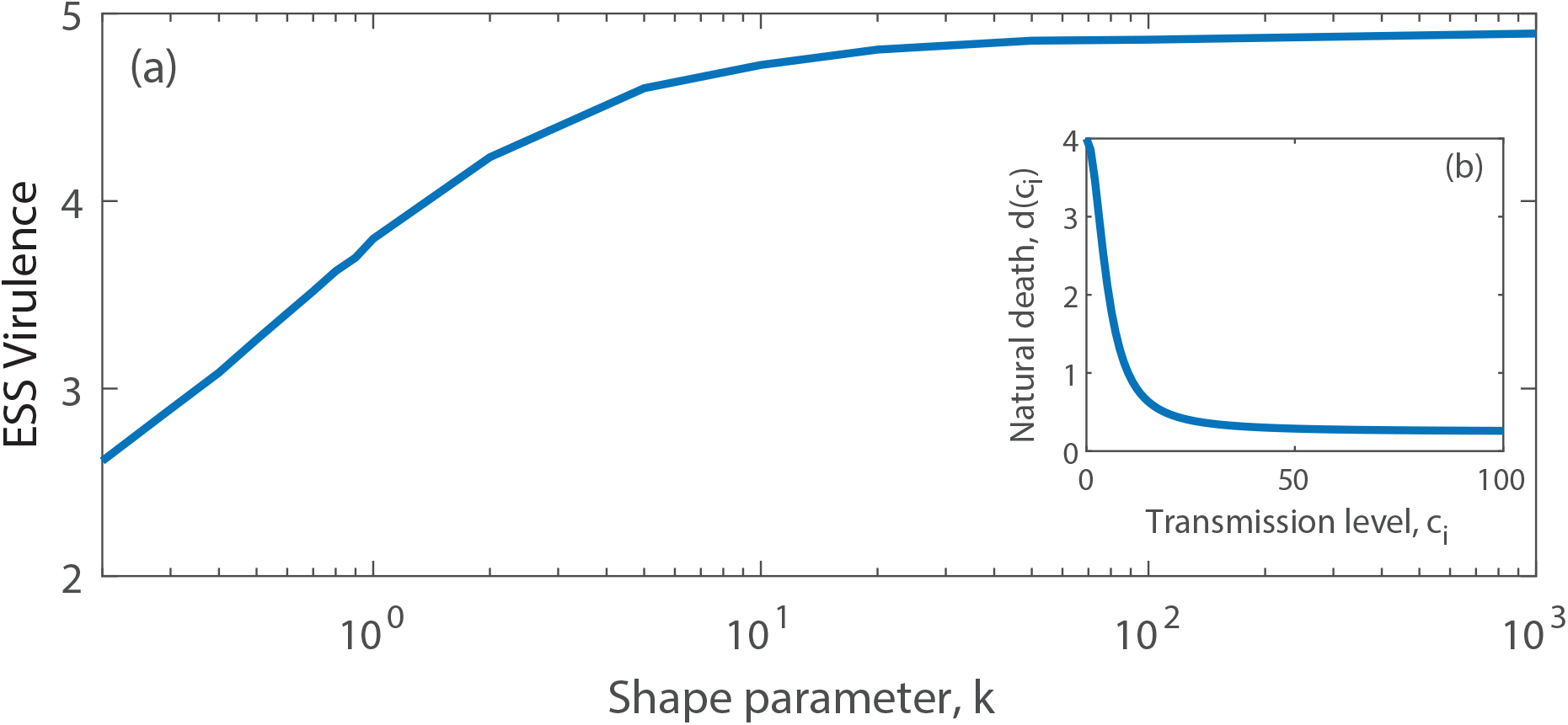
The evolution of virulence for the *SI* model when contacts are linked to host survival. (a) The evolved level of pathogen virulence, *α*^*∗*^, for different transmission distributions for the host (characterised by changes in *k*), and with rates of host natural death, *d*(*c*_*i*_), linked to host transmission level, *c*_*i*_. (b) The function *d*(*c*_*i*_) where the host death rate decreases with increases in host transmission level (increased connectivity). Results are obtained from deterministic simulations using parameters as in Figure 2 and the function 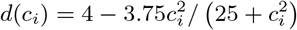.

Under superspreading hosts with high contact levels make the greatest contribution to transmission (see Figure S.3), and these hosts have a longer lifespan. In line with previous theory, when overall host lifespan increases, the parasite will reduce virulence [15, 18].

#### Superspreading due to variation in tolerant individuals

Variation in the transmission of infection often arises due to differences in tolerance to infection across the host population [39, 40, 52], as highlighted by the classic example for typhoid [53]. Here, superspreaders have high pathogen loads but are tolerant, showing few signs of disease, and therefore cause a disproportionate number of transmission events [7, 10, 11]. This implies that *h*(*c*_*i*_) is a decreasing function of *c*_*i*_ (Figure 4b). Here, we find that pathogen virulence increases as the transmission distribution for the host changes from homogeneous to superspreading (Figure 4a). Under superspreading, a greater proportion of infected hosts are tolerant since they have reduced mortality when infected (see Figure S.4). This reduces the cost of virulence for the pathogen and therefore drives an increase in virulence and transmission. Thus, a key result is that if tolerance underlies superspreading it will select for higher virulence.

**Figure 4:**
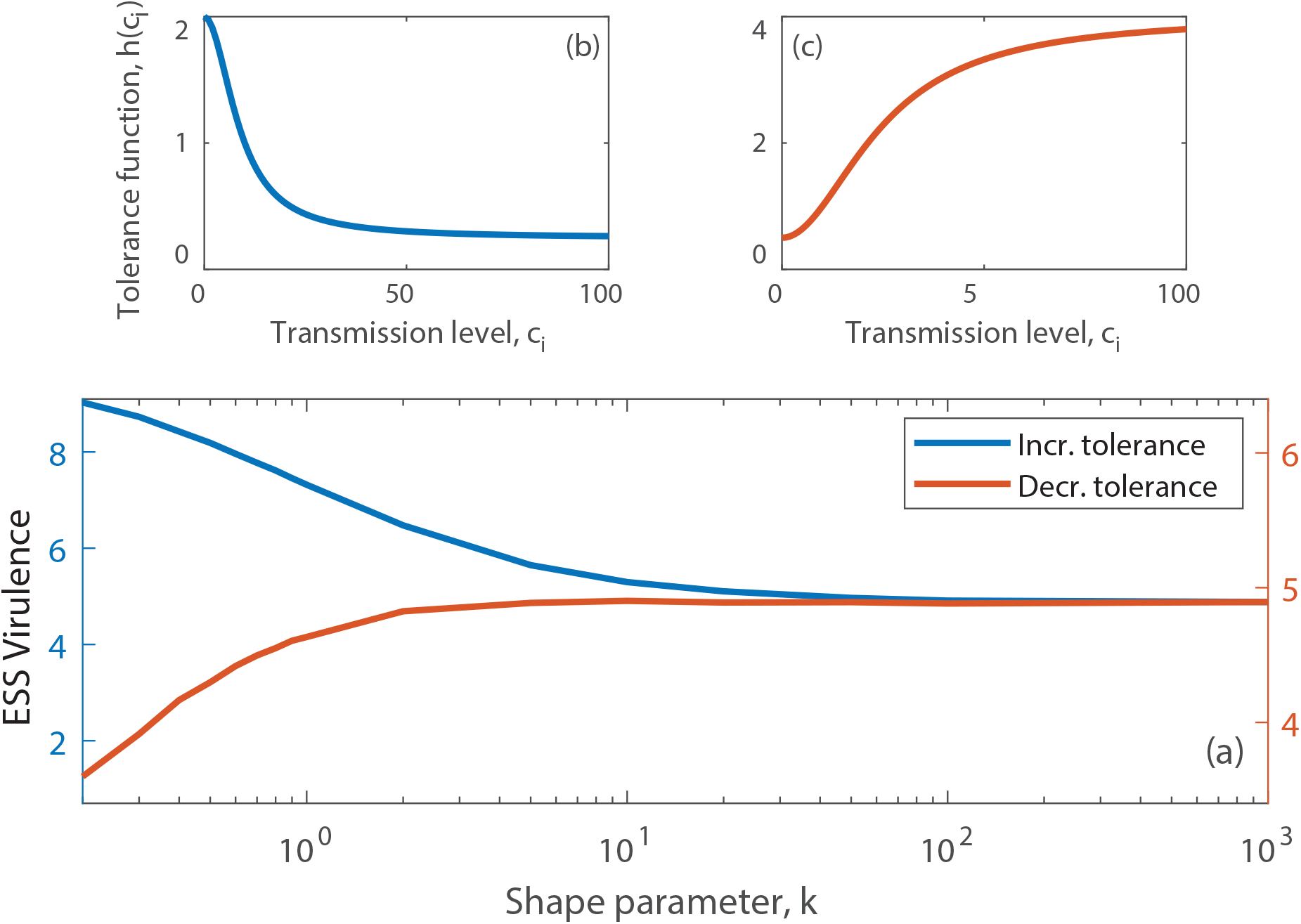
The evolution of virulence for the *SI* model when transmission is linked to tolerance or vulnerability. (a) The evolved level of pathogen virulence, *α*^*∗*^, for different transmission distributions for the host (characterised by changes in *k*), and with the function *h*(*c*_*i*_), linked to host transmission level, *c*_*i*_. (b) The function *h*(*c*_*i*_) where increases in *c*_*i*_ lead to increased tolerance (a decrease in *h*(*c*_*i*_)). (c) The function *h*(*c*_*i*_) where increases in *c*_*i*_ lead to increased vulnerability (a increase in *h*(*c*_*i*_)). Results are obtained from deterministic simulations using parameters as in Figure 2 and the functions (b) 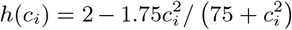 and (c) 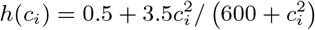.

While it may be reasonable to assume that tolerant superspreaders are more infective, this may not imply that they are also more susceptible to infection [8, 34]. Therefore, we repeat our analysis under the assumption that susceptibility is constant (at 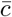) and therefore the transmission distribution for the host captures the variation in infectivity of the host and the link between highly infective hosts and tolerance. Under these assumptions we again find that pathogen virulence increases under a superspreading distribution (Figure S.5).

#### Superspreading due to variation in vulnerable individuals

Conversely, if increases in transmission levels for hosts arise due to increased host vulnerability that manifest as a reduction in host tolerance to infection [39, 40, 52], then *h*(*c*_*i*_) is an increasing function of *c*_*i*_ (Figure 4c). We assume vulnerable hosts will have high susceptibility, high infectivity, and increased disease-induced mortality. We find that the evolved level of pathogen virulence decreases as the transmission distribution of the host changes from a more homogeneous to a heterogeneous distribution (Figure 4a, Figure S.6). Under heterogeneous transmission, the hosts with high transmission (high *c*_*i*_) are more easily infected. These hosts have increased disease-induced mortality (high *h*(*c*_*i*_)), which increases the cost of virulence to the pathogen. To compensate, the pathogen evolves to reduce the component of virulence under its control (a reduction in *α*^*∗*^).

## Discussion

In order to understand the implications of superspreading on the evolution of pathogens we have developed a general mathematical framework that examines the evolution of pathogen virulence under a range of different distributions of infection transmission for the host and its relationship with other host and pathogen characteristics. Following established epidemiological theory, we characterise infection transmission for the host by a gamma distribution [11] and adjust the shape parameter to alter the transmission distribution from homogeneous to superspreading. In line with previous studies that consider classical *SI* model frameworks, when infection transmission for the host is independent of other host characteristics, we find that superspreading does not affect the evolutionarily stable level of virulence [32, 36], but it does have an important impact on the rate of evolution to the ESS [13, 14]. We show that this result extends to other model frameworks (*SIS, SIR, SIRS*) and holds when transmission is density-dependent or frequency-dependent. By considering stochastic simulations for the long-term evolution of virulence, our work highlights that superspreading can also lead to increased variation in virulence. If there is a link between the transmission level of the host and host mortality, we show that superspreading can have a range of impacts on the selection for virulence. In particular, superspreading may arise due to increased tolerance to infection or asymptomatic infection, such that the disease-induced death rate is reduced. In this case, selection for pathogen virulence increases as the transmission distribution for the host changes from homogeneous to superspreading. In contrast if high levels of transmission by the host are associated with a decrease in non-disease mortality or an increase in disease mortality, the evolved level of pathogen virulence will decrease as the transmission distribution for the host changes from homogeneous to heterogeneous. It is well known [11] that the transmission of many infectious diseases can be characterised by superspreading, and our work shows that this can have important implications for the evolution of pathogen virulence.

The evolutionary implications of superspreading can be understood through its impacts on the epidemiological dynamics. We found that a general consequence of superspreading is that the rate of evolution is reduced and the variation in levels of virulence is increased. These effects arise because as the transmission distribution for the host transitions from homogeneous to superspreading, there is an increase in the proportion of infected individuals that have high transmission levels and/or susceptible individuals that have low transmission. Lloyd-Smith et al. (2005) [11] showed that both theory and data indicated that superspreading leads to less frequent but more pronounced outbreaks of infection. The reduction in outbreak frequency emerges because the initial infection is likely to occur in a susceptible host with a low transmission level, and hence the infection may suffer stochastic extinction. The ability of new strains to invade an established, endemic strain has been shown to be linked to pathogen emergence [12–14]. Moreover, it has been shown that superspreading can reduce the chance of an emergence event and so would be expected to slow the rate of adaptation of a pathogen [13, 14]. In this study we examine the long-term evolutionary dynamics of the pathogen (we consider multiple emergence events). Under superspreading dynamics, a mutant pathogen strain is likely to emerge, on average, in an individual that has a low transmission level and is therefore more likely to suffer stochastic extinction. Thus, repeated mutation events may be required before the mutant strain is selected for, and this leads to a reduced rate of evolution towards the ESS under superspreading. One of our key results is that superspreading leads to increased variation around the optimal strain. This suggests that the adaptation to its new host of an emerging pathogen that shows high levels of superspreading, may generate and maintain multiple maladapted variants (while evolving to and subsequently around the optimal strategy), which could be more virulent than is optimal in the long-term. Our results that consider different distributions for infection transmission for the host have parallels with results from population genetics, where a reduced effective population size can increase the chance of drift [54]. Read and Keeling (2003) [55] showed that evolutionary change is slower and more variable for a spatial network, than for a comparable mean-field framework, since the network was influenced by the local epidemiology, that can lead to increased extinction of strain lineages, but also because the network models have fewer connections than the mean field framework [55]. The transmission distributions we consider all have the same mean level of connections, and the reduced rate of evolution and increased variability emerge due to the impact of superspreading on the epidemiological dynamics, where mutations arise in individuals with low transmission levels and so have a high chance of stochastic extinction (low chance of emergence [13, 14]).

A key result from our study is that the transmission distribution of the host can influence the evolution of virulence when transmission is associated with tolerance to the disease that a pathogen causes. It is likely that tolerant or asymptomatic individuals will, on average, cause more transmission events due to their lack of symptoms and could therefore drive superspreader events [7, 10, 11, 34]. Tolerance to pathogens is an important defence mechanism that reduces the harm that infection causes the host potentially lowering the mortality effect of infection [42, 52, 56]. Hosts that are more tolerant, live longer, leading to an increase in the infectious period and pathogen prevalence in the population and tolerance has been shown to select pathogens for higher replication rates, and therefore higher pathogen transmission and virulence in non-tolerant hosts [39, 40, 52, 57]. This explains our result that when hosts with high infectivity are more tolerant the pathogen will evolve higher virulence under superspreading. Similar mechanisms lead to selection for higher intrinsic virulence in response to imperfect vaccines that reduce the within host growth rate of the pathogen [30, 31] and so our work emphasises how these established theoretical results can be viewed in a new context of superspreading. Our result provides further support for surveillance and testing during infectious outbreaks, and for targeted treatment, since asymptomatic (tolerant) individuals may be responsible for an increase in the incidence of infection [11, 34, 58, 59]. Our study shows that they may also select for more virulent pathogen strains.

Previous studies that have examined how host contact rates may affect the evolution of virulence have generally assumed host contact to be independent of host survival [17, 19, 60]. Although high host contact levels typically increase the risk of infection, evidence from wildlife systems suggests that sociability has benefits [50, 51]. For example, social information can provide individuals with knowledge about resources or environmental conditions, which can reduce host mortality. Ravens, for instance, share information about the location of carcasses with conspecifics [61], and matriarchs of African elephant herds share information about danger and resources with the group [62]. Bonds et al. (2005) [32] showed that when an increase in contact rates can reduce non-disease mortality, the associated increase in host lifespan can indirectly select for lower pathogen virulence [32, 33]. This is in line with the general result that indicates that an increase in host lifespan can lead to the evolution of reduced virulence [18]. Our study builds on these insights by examining the evolution of virulence under different host contact distributions (with the same mean contact level) where individual hosts with high contact levels have a fitness benefit in terms of reduced natural mortality. We demonstrate that altering transmission distributions toward superspreading alters contact patterns within susceptible and infected populations, reshaping the epidemiological dynamics and leading to selection for lower pathogen virulence. While there are few natural examples directly illustrating how contact distribution affects virulence, studies in socially structured wildlife populations suggest that transmission dynamics in these systems mirror our models predictions. For example, superspreader events in vampire bat colonies play a crucial role in rabies transmission [63]. Superspreading bats are socially central and thus may benefit from better health via social grooming and foraging behaviours, which thus may prolong the infectious period and favour the evolution of lower virulence. Similar dynamics exist in tuberculosis spread in both African buffalo [64] and meerkats [65], where social support in highly connected (and thus superspreading) individuals may reduce stress and prolong infection. These cases suggest that in animal populations superspreading often occurs within social structures, creating correlations between transmission and fitness that aligns with our models assumptions. Together, these examples and our findings illustrate that social structure in animal populations can influence the evolution of virulence, providing real-world context for the implications of superspreading on pathogen evolution.

We present a general framework for assessing how superspreading influences the evolution of pathogen virulence. Our work emphasizes the importance of including epidemiological dynamics when determining the evolution of pathogen characteristics, since our evolutionary results are direct consequences of changes in these dynamics. Furthermore, our methods can be extended to assess the role of superspreader events under a wide range of scenarios. This could include exploring the impact of heterogeneous contact rates on the evolution of virulence in explicitly spatial settings, to tailor the model framework to specific infectious disease systems or to examine how the level of heterogeneity in pathogen transmission may itself evolve. The risk of epidemics in human, agricultural and wildlife systems is intensifying [3], and therefore our findings have important consequences for understanding the risks from rapidly evolving, superspreading, pathogens.

## Supporting information

Supplementary material, used for link to the file on the preprint site.

## Acknowledgements

X.O. and A.W. were supported by the BBSRC EEID research grant BB/V00378X/1. M.B., G.R.N., and P.S.W. were supported by NSF-DEB-2011109 EEID research grant. C.M.S.-R. gratefully acknowledges funding from the Miller Institute for Basic Research in Science of UC Berkeley via a Miller Research Fellowship. The funders had no role in study design, data collection and analysis, decision to publish, or preparation of the manuscript.

## Author Contributions

X.O., A.W. and M.B. designed the study. X.O. and A.W. performed the theoretical and computational analysis. A.W. and X.O. wrote the initial draft of the manuscript with key contributions from all authors. All authors approved the final version.

## Author Declaration

The authors have declared that no competing interests exist.

## Data Availability Statement

Please use the following link to access example code in MAT-LAB2023b:

https://github.com/XanderONeill/Superspreading_and_the_evolution_of_virulence

## Supplementary Material

The supplementary material contains the following sections: S.1 Adaptive dynamics analysis when transmission is independent of other host characteristics, S.2 Trade-off function, S.3 Other deterministic and stochastic model simulations.

