## Supplementary material, used for link to the file on the preprint site. for "Superspreading and the evolution of virulence"

Supplementary Information
Superspreading and the evolution of virulence

### S.1 Adaptive dynamics analysis when transmission is independent of other host characteristics

When transmission is independent of other host characteristics the host contribution to virulence $h(c_i) = 1$ and host natural death rate $d(c_i) = d = 1$ are constant for all host types. The $SI$ version of the model (Equations 1 in the main text) becomes:

$$\frac{dS_i}{dt} = N\left(b - qN\right)p_i - \beta c_i S_i \sum_j c_j I_j - dS_i,$$
$$\frac{dI_i}{dt} = \beta c_i S_i \sum_j c_j I_j - \left(d + \alpha\right) I_i. \tag{S.1}$$

We use adaptive dynamics to explore the effect of transmission structure of the host on the evolution of virulence, $\alpha$. Adaptive dynamics assumes a separation of epidemiological and evolutionary time scales such that the epidemiological dynamics have reached an endemic steady state before a new mutation is considered. When a mutation occurs a mutant strain, with small phenotypic variation from the resident strain, is rare and attempts to invade the resident system at its endemic steady state [1, 2]. To assess how virulence will evolve we derive the fitness function for a mutant strain of infection and determine conditions that allow the mutant to invade the resident population.

We consider a mutant strain of the pathogen with parameters $\alpha_M$ and $\beta_M$ that attempts to invade a resident pathogen strain with parameters $\alpha_R$ and $\beta_R$. The fitness, $r_M$, of the mutant strain can be determined from the spectral radius (largest eigenvalue) of the mutant sub-matrix of the Jacobian matrix evaluated at the mutant-free, endemic, steady state [3]. To provide an explicit description of how the fitness function is calculated let us consider $n_c = 2$ host types, with transmission levels $c_1$ and $c_2$ and probabilities of being born with these transmission levels $p_1$ and $p_2$, respectively, and where $p_1 + p_2 = 1$ and $c_1 p_1 + c_2 p_2 = \bar{c}$. The resident and mutant dynamics are as follows:

$$\begin{aligned}
\frac{dS_1}{dt} &= N\left(b-qN\right)p_1 - \beta c_1 S_1 (c_1 I_{1R} + c_2 I_{2R} + c_1 I_{1M} + c_2 I_{2M}) - dS_1,\\
\frac{dS_2}{dt} &= N\left(b-qN\right)p_2 - \beta c_2 S_2 (c_1 I_{1R} + c_2 I_{2R} + c_1 I_{1M} + c_2 I_{2M}) - dS_2,\\
\frac{dI_{1R}}{dt} &= \beta_R c_1 S_1 (c_1 I_{1R} + c_2 I_{2R}) - \left(d+\alpha_R\right) I_{1R},\\
\frac{dI_{2R}}{dt} &= \beta_R c_2 S_2 (c_1 I_{1R} + c_2 I_{2R}) - \left(d+\alpha_R\right) I_{2R},\\
\frac{dI_{1M}}{dt} &= \beta_M c_1 S_1 (c_1 I_{1M} + c_2 I_{2M}) - \left(d+\alpha_M\right) I_{1M},\\
\frac{dI_{2M}}{dt} &= \beta_M c_2 S_2 (c_1 I_{1M} + c_2 I_{2M}) - \left(d+\alpha_M\right) I_{2M},
\end{aligned} \tag{S.2}$$

where $I_{1R}, I_{2R}$ denote the density of hosts infected with the resident pathogen strain and with transmission levels $c_1$ and $c_2$, respectively, and $I_{1M}, I_{2M}$ denote the density of hosts infected with the mutant pathogen strain and with transmission levels $c_1$ and $c_2$, respectively. The mutant strain sub-matrix of the Jacobian, $J_{mut}$, evaluated at the mutant-free, endemic, steady state is as follows:

$$J_{mut} = \begin{pmatrix} \beta_M (c_1)^2 S_1 - \alpha_M - d & \beta_M c_1 c_2 S_1 \\ \beta_M c_1 c_2 S_2 & \beta_M (c_2)^2 S_2 - \alpha_M - d \end{pmatrix}, \tag{S.3}$$

where here $S_1$ and $S_2$ represent the susceptible density at the mutant-free, endemic, steady state. The eigenvalues of this matrix are given by $\lambda_{1,2}$ as follows:

$$\lambda_1 = -d - \alpha_M, \qquad \lambda_2 = \beta_M (c_1)^2 S_1 + \beta_M (c_2)^2 S_2 - \alpha_M - d.$$

As $\lambda_2 > \lambda_1$, the spectral radius, and therefore fitness expression ($r_M$) for the mutant pathogen strain, is represented by $\lambda_2$.

This method for determining the mutant pathogen fitness expression can extend to cases with more than two transmission levels ($n_c > 2$). The fitness of the mutant pathogen strain, $r_M$, is given by the following expression:

$$r_M = f(\alpha_M) \sum_{i=1}^{n_c} c_i^2 S_i - (\alpha_M + d). \tag{S.4}$$

Here $S_i$ represents the steady state density of susceptible host type $i$ for the resident and $f(\alpha_M) = \beta_M$. By definition, the fitness, $r_R$, of the resident population is zero, and therefore

$$\sum_{i=1}^{n_c} c_i^2 S_i = \frac{\alpha_R + d}{f(\alpha_R)}. \tag{S.5}$$

Using Equation (S.5) we can rewrite Equation (S.4) to show that the fitness of the mutant strain is positive, $r_M > 0$, if the following condition is satisfied:

$$\frac{f(\alpha_M)}{(\alpha_M + d)} > \frac{f(\alpha_R)}{(\alpha_R + d)}. \tag{S.6}$$

Therefore, any invading mutant strain that satisfies Equation (S.6) will replace the resident strain and the pathogen evolves a level of virulence, $\alpha^*$, that maximizes $f(\alpha)/(\alpha + d)$, which is the optimal strategy [4]. The evolutionary singular strategy therefore satisfies the following condition:

$$f'(\alpha^*) = \frac{f(\alpha^*)}{\alpha^* + d}. \tag{S.7}$$

This is independent of the transmission level of the host and so when $h(c_i) = 1$ and $d(c_i) = d$ pathogen virulence will evolve to an evolutionarily singular strategy (ESS) at $\alpha^*$ for all transmission distributions of the host.

#### S.1.1 Other model frameworks

Under the same assumptions, other model structures ($SIS, SIR, SIRS$) also lead to the finding that $\alpha^*$ is independent of the transmission level of the host. We specifically show the analysis for the $SIRS$ model framework as this encompasses the other model frameworks. The modification of Equations (1) to represent an $SIRS$ epidemiological framework is as follows:

$$\begin{aligned}
\frac{dS_i}{dt} &= N\left(b - qN\right)p_i - \beta c_i S_i \sum_j c_j I_j - dS_i + \nu R_i, \\
\frac{dI_i}{dt} &= \beta c_i S_i \sum_j c_j I_j - \left(d + \alpha + \gamma\right) I_i, \\
\frac{dR_i}{dt} &= \gamma I_i - dR_i - \nu R_i,
\end{aligned} \tag{S.8}$$

where $\gamma$ is the rate of recovery of infected individuals to the immune class, $R$, and $\nu$ is the rate at which individuals lose immunity and become susceptible once again. We again consider a mutant strain of the pathogen with parameters $\alpha_M$ and $\beta_M$ that attempts to invade a resident pathogen strain with parameters $\alpha_R$ and $\beta_R$. The fitness, $r_M$, of the mutant strain can be determined as follows:

$$r_M = f(\alpha_M)\sum_{i=1}^{n_c} c_i^2 S_i - (\alpha_M + d + \gamma), \tag{S.9}$$

where $S_i$ represents the steady state density of susceptible host type $i$ for the resident. By definition, the fitness, $r_R$, of the resident population is zero, and therefore

$$\sum_{i=1}^{n_c} c_i^2 S_i = \frac{\alpha_R + d + \gamma}{f(\alpha_R)}. \tag{S.10}$$

Using Equation (S.10) we can rewrite Equation (S.9) to show that $r_M > 0$ if the following expression holds.

$$\frac{f(\alpha_M)}{\alpha_M + d + \gamma)} > \frac{f(\alpha_R)}{(\alpha_R + d + \gamma)}. \tag{S.11}$$

Therefore, any invading mutant strain that satisfies Equation (S.11) will replace the resident strain and the pathogen evolves a level of virulence, $\alpha^*$, that maximizes $f(\alpha)/(\alpha + d + \gamma)$, which is the optimal strategy. Again, this is independent of the transmission level of the host and so pathogen virulence will evolve to an ESS at $\alpha^*$ for all transmission distributions of the host.

### S.1.2 Frequency-dependent transmission

The results we have considered so far assume density-dependent infection transmission but our findings extend to frameworks that consider frequency-dependent infection transmission. Again, we show the analysis for the $SIRS$ model framework as this encompasses the other model frameworks. The $SIRS$ epidemiological framework with frequency-dependent transmission is as follows:

$$\begin{aligned}
\frac{dS_i}{dt} &= N(b - qN)p_i - \frac{\beta}{N} c_i S_i \sum_j c_j I_j - dS_i + \nu R_i, \\
\frac{dI_i}{dt} &= \frac{\beta}{N} c_i S_i \sum_j c_j I_j - (d + \alpha + \gamma) I_i, \\
\frac{dR_i}{dt} &= \gamma I_i - dR_i - \nu R_i.
\end{aligned} \tag{S.12}$$

Following the methods outlined previously, the fitness, $r_M$, of the mutant strain can be determined as follows:

$$r_M = \frac{f(\alpha_M)}{N} \sum_{i=1}^{n_c} c_i^2 S_i - (\alpha_M + d + \gamma), \tag{S.13}$$

where $S_i$ represents the steady state density of susceptible host type $i$ for the resident and $N$ is the total population density at the resident endemic steady state (in the absence of the mutant). By definition, the fitness, $r_R$, of the resident population is zero, and therefore

$$\frac{1}{N}\sum_{i=1}^{n_c} c_i^2 S_i = \frac{\alpha_R + d + \gamma}{f(\alpha_R)}. \tag{S.14}$$

Using Equation (S.14) we can rewrite Equation (S.13) to show that $r_M > 0$ if the following expression holds.

$$\frac{f(\alpha_M)}{(\alpha_M + d + \gamma)} > \frac{f(\alpha_R)}{(\alpha_R + d + \gamma)}. \tag{S.15}$$

Therefore, any invading mutant strain that satisfies Equation (S.15) will replace the resident strain and the pathogen evolves a level of virulence, $\alpha^*$, that maximizes $f(\alpha)/(\alpha + d + \gamma)$, which is the optimal strategy. Again, this is independent of the transmission level of the host and so in the model framework with frequency-dependent transmission pathogen virulence will evolve to an ESS at $\alpha^*$ for all transmission distributions of the host.

### S.2 Trade-off function

The trade-off function represented by Equation (3) in the main text is shown in Figure S.1.

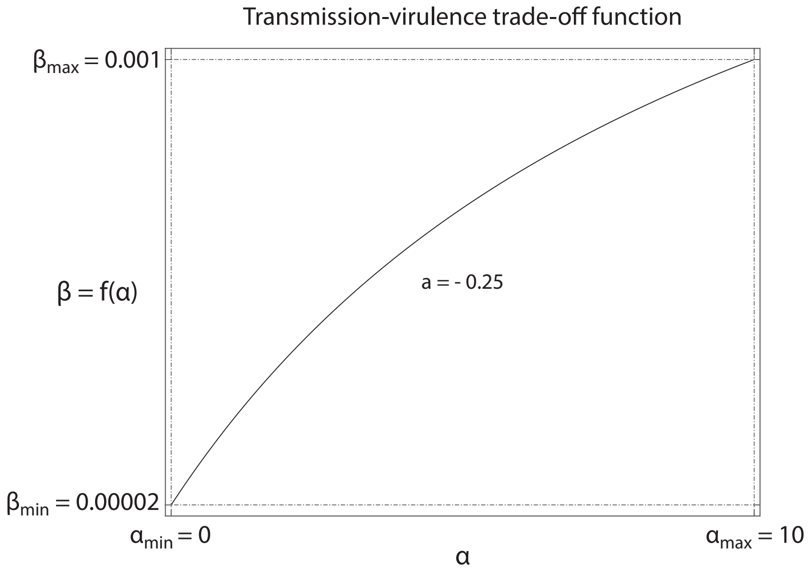


**Figure S.1:** Graphical representation of the transmission-virulence trade-off function (Equation 3 in the main text).

### S.3 Deterministic and Stochastic Simulations

We present further deterministic and stochastic simulations that correspond to the figures in the main text.

Figure S.2 relates to Figure 2.

Figure S.3 relates to Figure 3.

Figure S.4, S.5 and S.6 relate to Figure 4.

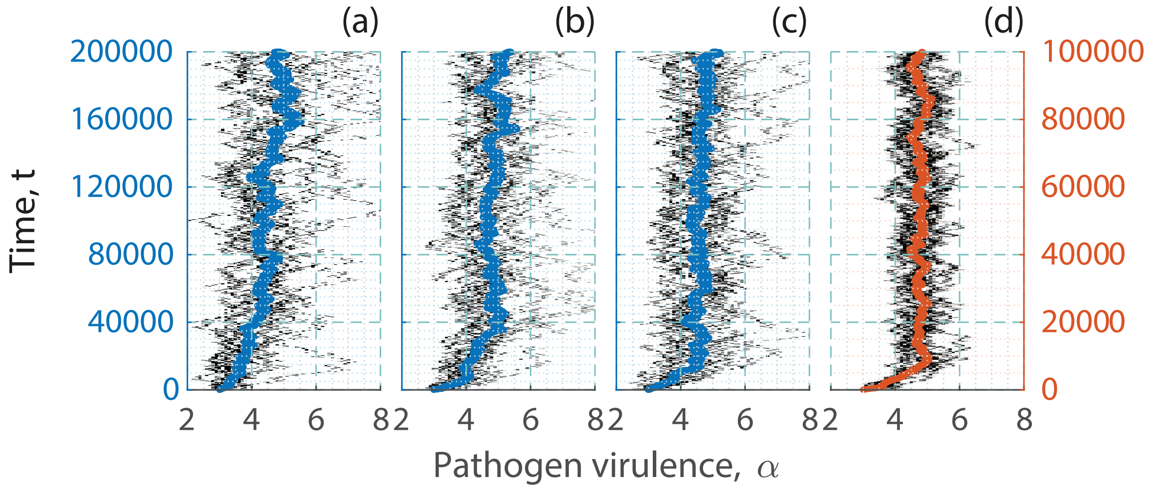


**Figure S.2:** Stochastic simulations of the evolution of pathogen virulence over time when infection transmission for the host is independent of other host characteristics. In (a)-(c) hosts have a super-spreader/heterogeneous distribution ($k = 0.2$) where in (a) transmission depends on susceptibility and infectivity ($\beta c_i c_j$), in (b) transmission depends on infectivity ($\beta \bar{c} c_j$) and in (c) transmission depends on susceptibility ($\beta c_i \bar{c}$). In (d) hosts have a more homogeneous distribution ($k = 10$) and transmission depends on susceptibility and infectivity ($\beta c_i c_j$). Note, the vertical axis is different for (a) to (c) compared to (d). Parameters are the same as in Figure 2.

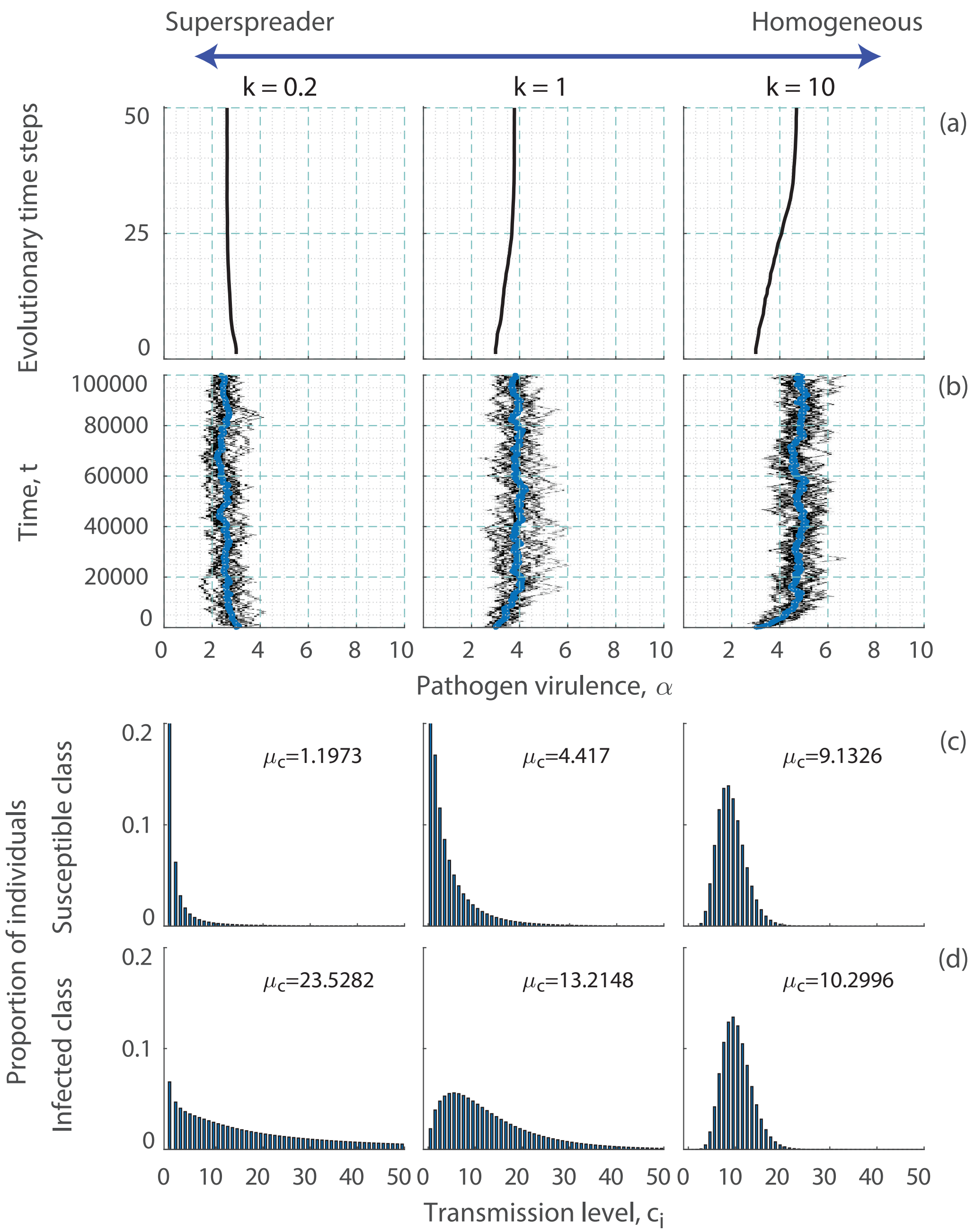


**Figure S.3:** The evolution of virulence when contacts are linked to host survival. In (a) and (b) we show the evolution of pathogen virulence over time under different transmission distributions. In (a) we show the deterministic simulations and (b) we show the stochastic simulations. In (c) we show the proportion of susceptible individuals in each transmission class, $c_i$ and (d) the proportion of infected individuals in each transmission class. All proportions are shown at the evolutionary stable level of pathogen virulence, $\alpha^*$, in the deterministic simulations. The mean level of transmission, $\mu_c$, is also shown for each distribution. The simulations are for a function $(d(c_i) = 4 - 3.75c_i^2/\left(25 + c_i^2\right))$ that shows a decrease in natural mortality for hosts that have high transmission, as in Figure 3. Other parameters are taken from Figure 2.

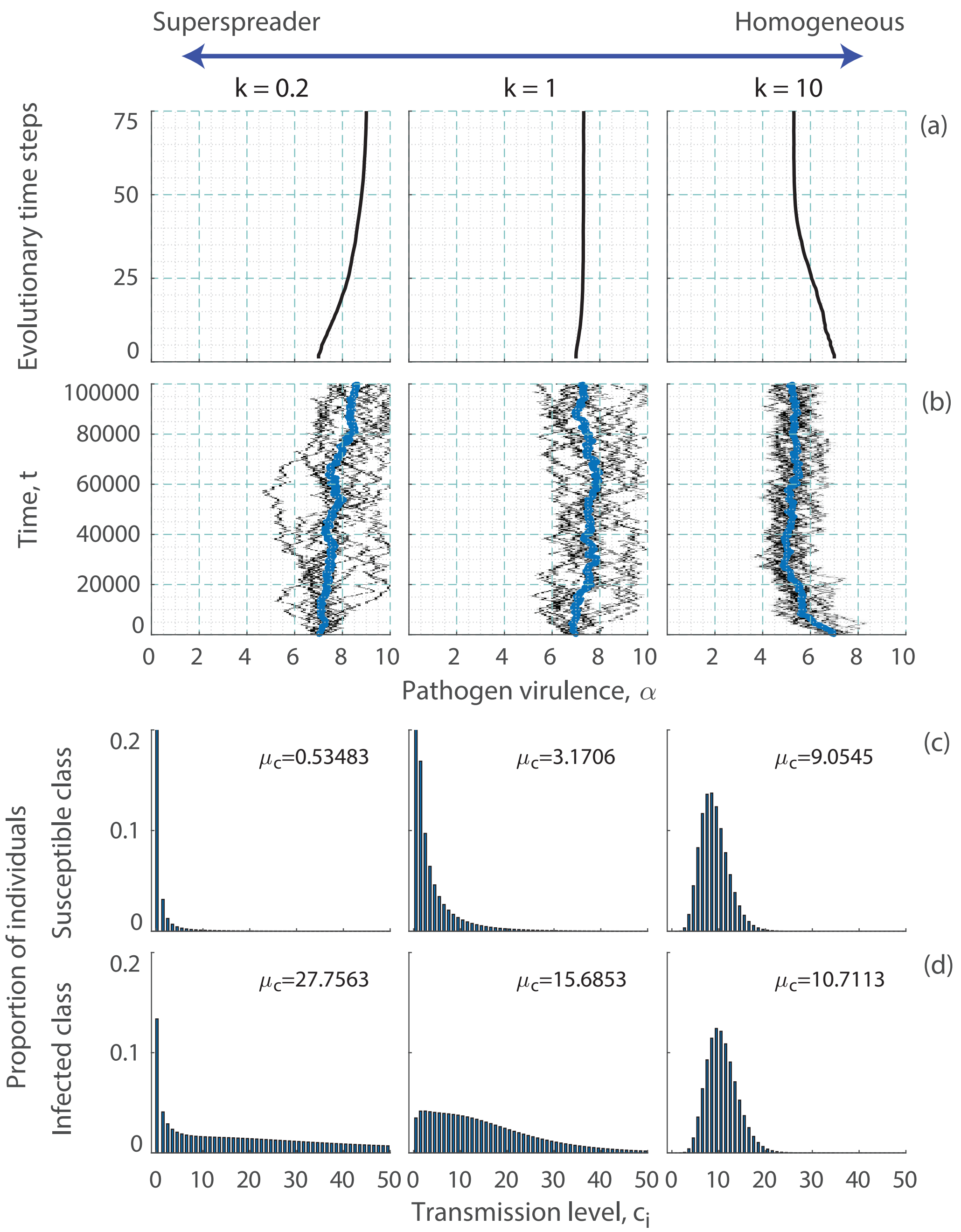


**Figure S.4:** The evolution of virulence when infection transmission for the host is linked to tolerance. In (a) and (b) we show the evolution of pathogen virulence over time under different transmission distributions. In (a) we show the deterministic simulations and (b) we show the stochastic simulations. In (c) we show the proportion of susceptible individuals in each transmission class, $c_i$ and (d) the proportion of infected individuals in each transmission class. All proportions are shown at the evolutionary stable level of pathogen virulence, $\alpha^*$, in the deterministic simulations. The mean level of transmission, $\mu_c$, is also shown for each distribution. The simulations are for a function $\left(h(c_i) = 2 - 1.75c_i^2/\left(75 + c_i^2\right)\right)$ that shows an increase in tolerance for hosts that have high transmission, as in Figure 4. Other parameters are taken from Figure 2.

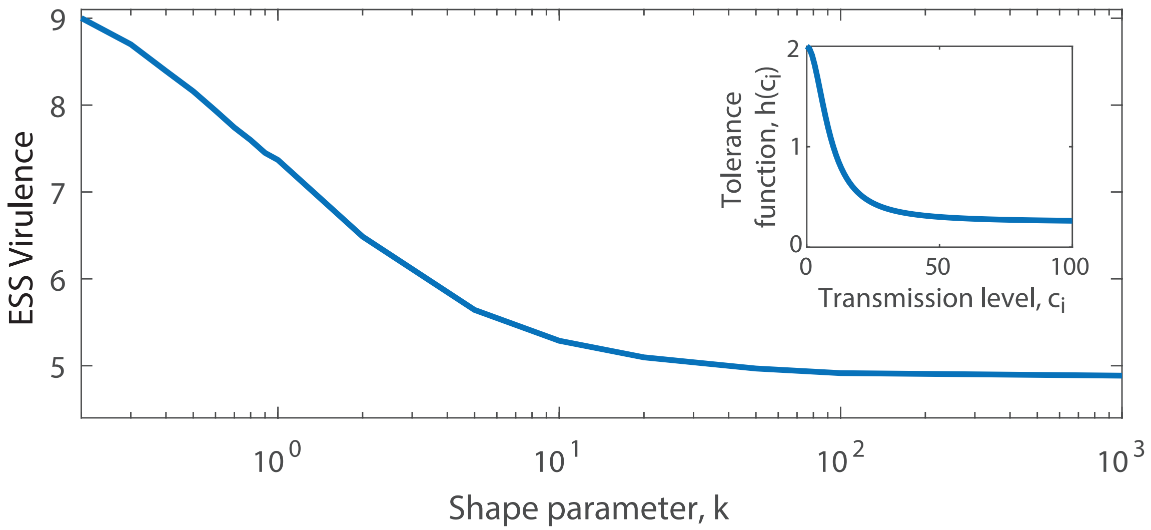


**Figure S.5:** The evolution of virulence when infectivity is linked to tolerance (increases in infectivity lead to increased tolerance) and when susceptibility is constant (at $\bar{c}$) for all hosts. The evolved level of pathogen virulence, $\alpha^*$, is shown for different transmission distributions (characterised by changes in $k$). Results are obtained from deterministic simulations using parameters as in Figure 2 and the function $(h(c_i) = 2 - 1.75c_i^2/\left(75 + c_i^2\right))$.

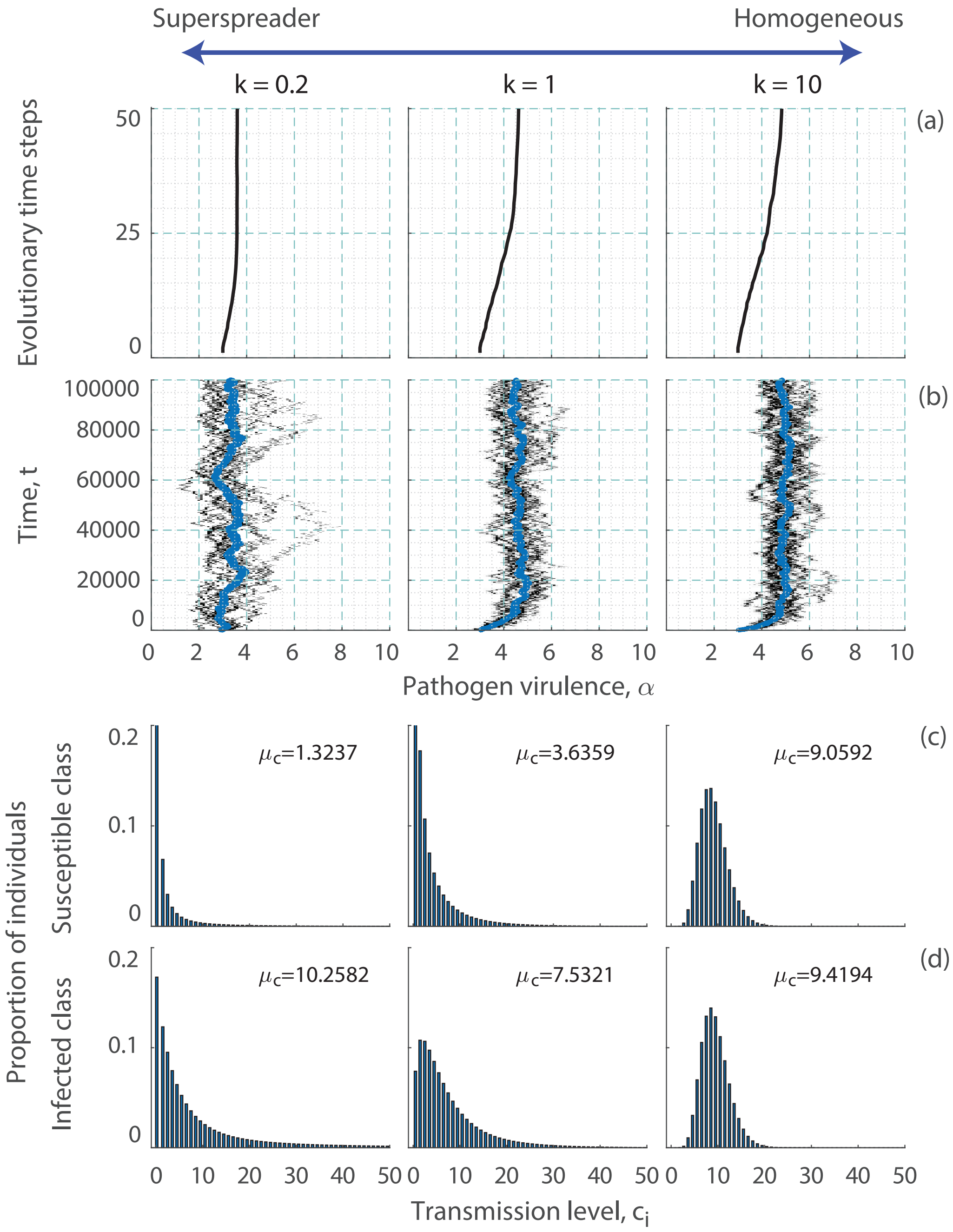


**Figure S.6:** The evolution of virulence when transmission is linked to vulnerability. In (a) and (b) we show the evolution of pathogen virulence over time under different transmission distributions. In (a) we show the deterministic simulations and (b) we show the stochastic simulations. In (c) we show the proportion of susceptible individuals in each transmission class, $c_i$ and (d) the proportion of infected individuals in each transmission class. All proportions are shown at the evolutionary stable level of pathogen virulence, $\alpha^*$, in the deterministic simulations. The mean level of transmission, $\mu_c$, is also shown for each distribution. The simulations are for a function $(h(c_i) = 0.5 + 3.5c_i^2/\left(600 + c_i^2\right))$ that shows an increase in vulnerability for hosts that have high transmission, as in Figure 4. Other parameters are taken from Figure 2.
